# Visually-biased perception in cochlear implant users: a study of the McGurk and sound-induced flash illusions

**DOI:** 10.1101/2021.04.09.21254577

**Authors:** Iliza M. Butera, Ryan A. Stevenson, René H. Gifford, Mark T. Wallace

## Abstract

The reduction in spectral resolution by cochlear implants oftentimes requires complementary visual speech cues to aid in understanding. Despite substantial clinical characterization of auditory-only speech outcome measures, relatively little is known about the audiovisual integrative abilities that most cochlear implant (CI) users rely on for daily speech comprehension. In this study, we tested audiovisual integration in 63 CI users and 69 normal-hearing (NH) controls using the McGurk and sound-induced flash illusions. This study is the largest to-date measuring the McGurk effect in this population and the first to test the sound-induced flash illusion. When presented with conflicting audiovisual speech stimuli (i.e., the phoneme “ba” dubbed onto the viseme “ga”), we found that 55 CI users (87%) reported a fused percept of “da” or “tha” on at least one trial. However, overall, we found that CI users experienced the McGurk effect less often than controls—a result that was concordant with results with the sound-induced flash illusion where the pairing of a single circle flashing on the screen with multiple beeps resulted in fewer illusory flashes for CI users. While illusion perception in these two tasks appears to be uncorrelated among CI users, we identified a negative correlation in the NH group. Because neither illusion appears to provide further explanation of variability in CI outcome measures, further research is needed to determine how these findings relate to CI users’ speech understanding, particularly in ecological listening conditions that are naturally multisensory.

## INTRODUCTION

Naturalistic oral communication is typically an audiovisual (AV) experience that reveals striking perceptual benefits in speech comprehension when visual cues are present (Sumby and Pollack 1954). In contrast, for cochlear implant (CI) users talking on the phone is a difficult, and oftentimes insurmountable, challenge—one that requires parsing auditory-only speech signals that are conveyed by the CI at greatly reduced spectral resolution. Fortunately, visual input can improve listening thresholds (Barone and Deguine, 2011; Barone et al., 2010; Grant and Seitz, 2000), and for many CI users, audiovisual conversations are a necessity.

Audiovisual integration is the process of filtering and combining information from the different senses, like auditory speech and visual articulations, in order to create a more accurate percept than either sense could on its own. Each sensory modality encodes complementary information that is increasingly useful when the saliency/interpretability of the individual signals is relatively low. While typical, acoustic listeners may only experience the benefits of audiovisual integration in, for example, a very noisy restaurant, electrical hearing through a CI may in itself introduce enough “noise” to necessitate the integration of visual cues as a critical compensatory strategy. In effect, many CI users rely on audiovisual integration to successfully understand speech in most listening environments, not just particularly noisy ones. As a result, visual orofacial articulations play a crucial role in verbal communication both before and after cochlear implantation, and in order to fully describe aural speech recovery following implant surgery, characterization of both unisensory and multisensory processing is necessary.

Despite a daily reliance on the multisensory integration of speech signals, audiovisual testing is not a part of routine audiological exams. Furthermore, as a clinical population, there are substantial individual differences (e.g., onset of hearing loss, etiology, duration of deafness, and age of implantation) that contribute to variable auditory outcome measures (Blamey et al. 2013) and may also have cascading effects on AV integration.

Arguably, the index of multisensory processing that we know the most about in CI users is the McGurk illusion (McGurk and MacDonald 1976). First described over 40 years ago, this illusion results from hearing a bilabial syllable (e.g., “ba” or “pa”) while seeing the visually-ambiguous articulation of a velar syllable (“ga” or “ka”). Together, these elicit a novel, fused percept such as “da”, “tha”, or “ta”. This effect is both robust and persistent for individuals who experience it, although it is well documented that a subset of individuals fail to perceive the illusion (Mallick, Magnotti, and Beauchamp 2015). Many studies of CI users have focused on the responses of these “non-perceivers,” because it provides insight into sensory biases in the absence of fused percepts.

One very consistent finding in all studies evaluating the McGurk illusion in CI users is that those who do not perceive the illusion are biased toward the visual component (see Table 1 for brief summaries and references). This finding is in contrast to NH controls who typically report the auditory component when not experiencing the illusion (Massaro, Thompson, and Barron 1986; McGurk and MacDonald 1976). This discrepancy makes intuitive sense, because many CI users struggle to discriminate auditory-only syllables, and may be perceptually “weighting” visual information more highly in order to improve AV estimates (Huyse, Berthommier, and Leybaert 2013). Indeed, adding noise or otherwise altering the saliency of one sensory modality is well-known to effectively and rapidly alter sensory weights in typical populations (Ernst and Bülthoff 2004).

**TABLE 1.**
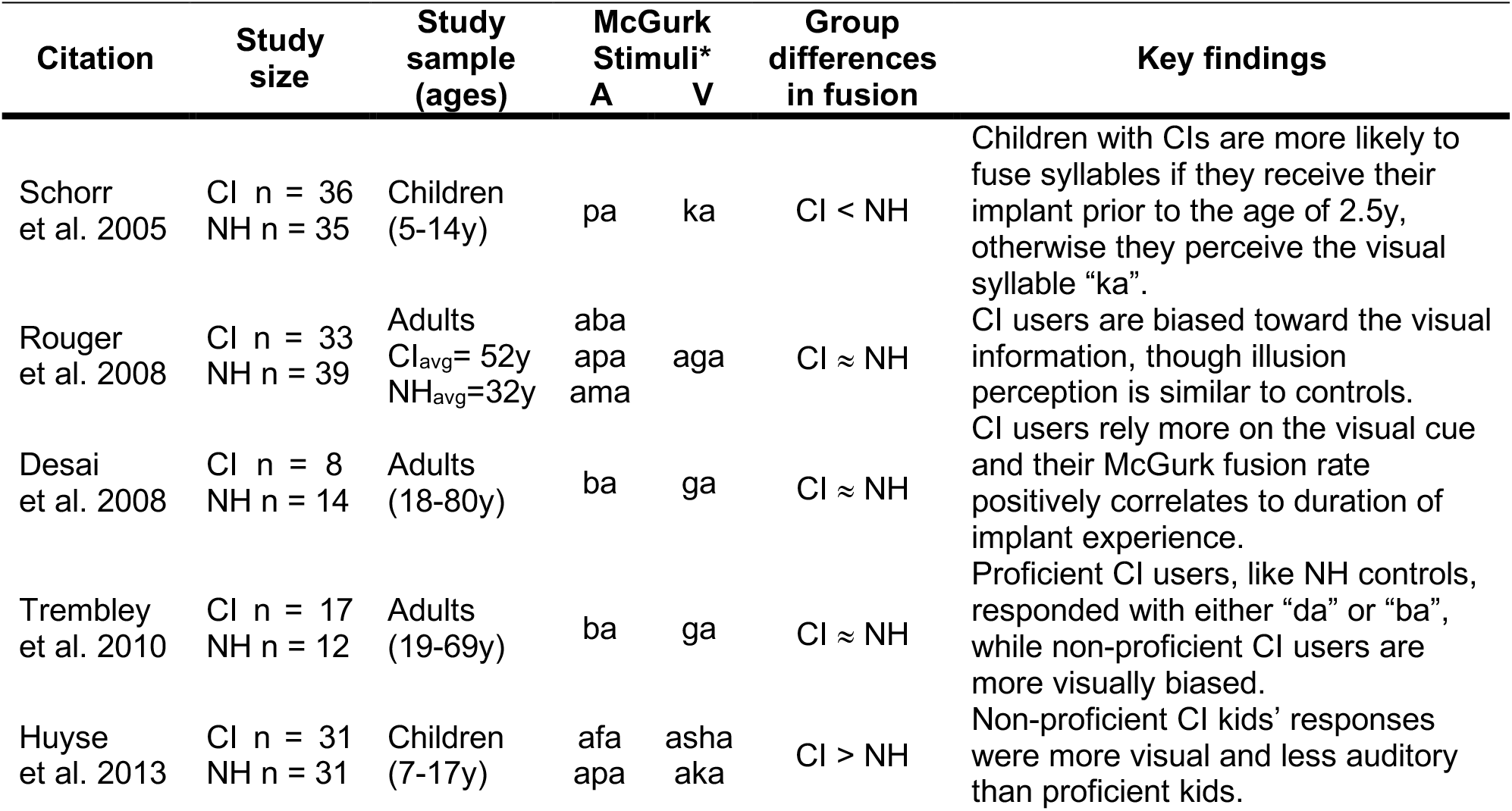
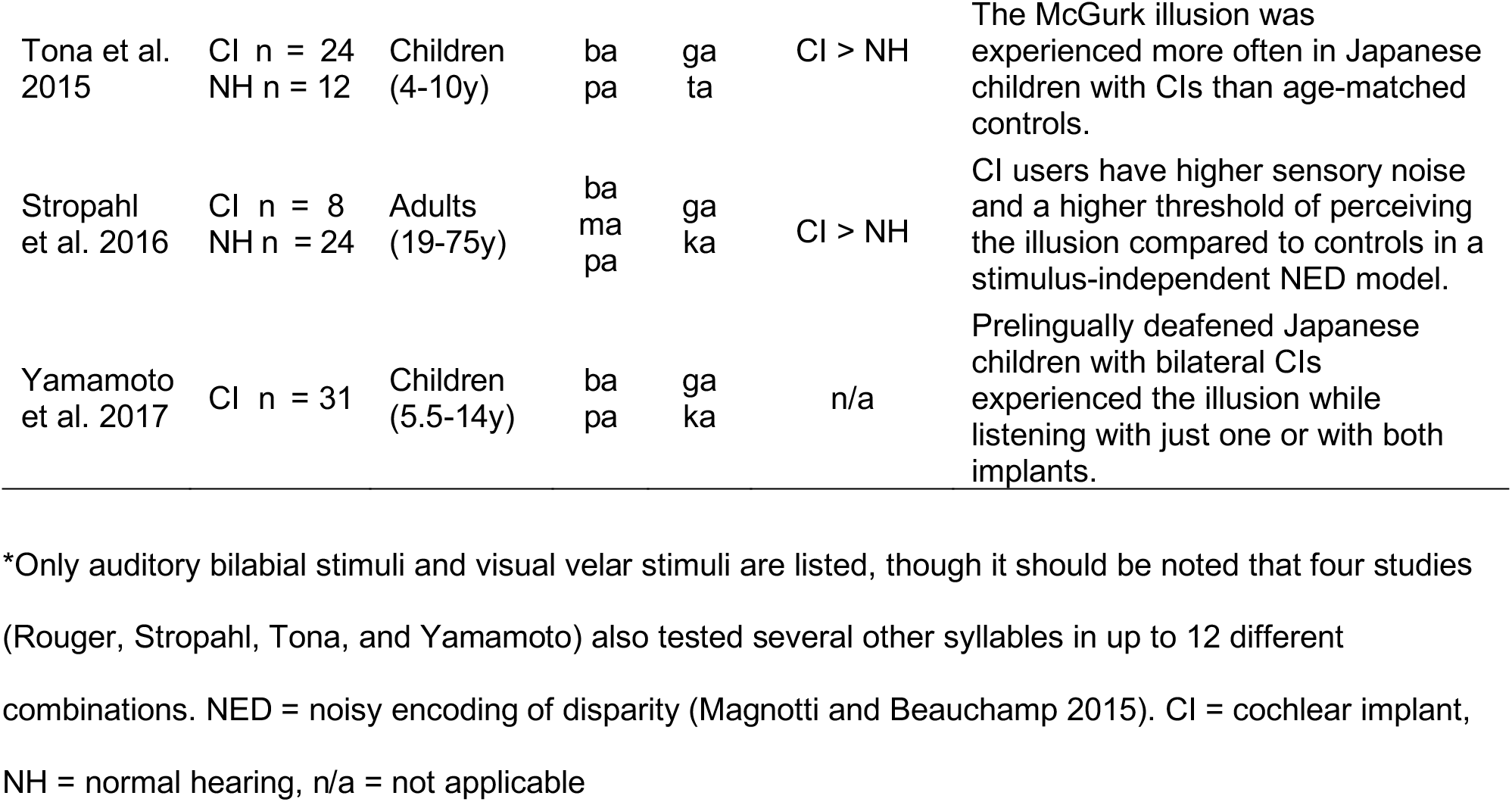
McGurk studies in the literature. All studies report a visual bias in CI users, while Schorr, Desai, and Tremblay also found various correlations to clinical measures.

Despite this consensus in the literature regarding visual bias among CI users, how their rate of fusion (e.g., “da” percepts) compares to NH controls is less apparent. Some studies indicate similar reports of fused percepts in CI users (Huyse et al. 2013; Rouger et al. 2008; Tremblay et al. 2010), while others find both lower (Schorr et al. 2005) and higher (Desai, Stickney, and Zeng 2008; Stropahl, Schellhardt, and Debener 2017; Tona et al. 2015) reports of McGurk percepts in CI users than controls (Table 1). Given that auditory saliency is lower in CI users (i.e., less reliable), direct comparisons to NH individuals requires further calculations and may account for some discrepancies in the literature. Take, for example, an auditory error in unisensory trials like mistaking the sound of “ba” for “da”. Because this response could also mimic a McGurk percept in the incongruent trials, an adjustment is necessary to better quantify illusion perception *per se* (Grant, Walden, and Seitz 1998; Stevenson, Zemtsov, and Wallace 2012). This need may be particularly true for patient populations with hearing impairments. Desai and colleagues, for instance, found much higher fusion response rates in CI users (60% v. 20%) that were actually not significantly different after applying an error correction (2008). Further investigations are needed to address this issue, particularly in larger sample sizes that also capture the clinical diversity of CI users today.

In addition to this outstanding question of whether McGurk perception truly differs between CI and NH listeners, it is also unknown how these results directly relate to other metrics of AV integration. The sound-induced flash illusion—sometimes referred to as the flashbeep or double flash illusion—was discovered more recently and involves simple, non-speech stimuli (Shams, Kamitani, and Shimojo 2000). Unlike the McGurk effect, the sound-induced flash illusion, or SIFI, asks participants to ignore what they hear and only report what they saw. Specifically, participants are asked to count the number of rings that rapidly flash on a screen while they also hear a varying number of beeps. The greater the number of beeps, the more likely that multiple, illusory flashes are perceived when only a single flash is presented. Given that deafness has been linked to perceptual enhancements in the visual periphery (Bavelier, Dye, and Hauser 2006) and this illusion is perceived more strongly in the parafoveal visual fields of typical listeners (Shams, Kamitani, and Shimojo 2002), employing this task in CI users may provide new context for underlying differences in audiovisual integration extending beyond speech stimuli. We hypothesized that the known visual biases in CI users would lead to more veridical visual perceptions, which would correspond to a *decreased* likelihood of perceiving the non-existent flashes in a SIFI task or the fused “da” percept in a McGurk task (see Fig. 1a schematic). Furthermore, because low McGurk fusion is effectively a metric of visual bias for CI users, we predicted a positive correlation between these two tasks; that is, when simple visual percepts are more easily biased by sound, we predicted a higher rate of fusion of auditory and visual speech signals leading to CI users perceiving the McGurk illusion (Fig. 1b). By the same logic, lower McGurk fusion in NH controls typically corresponds to an auditory-dominant percept, which could, in turn, influence a greater number of perceived flashes in the SIFI experiment (i.e., result in a negative correlation between the two tasks).

**FIGURE 1.**
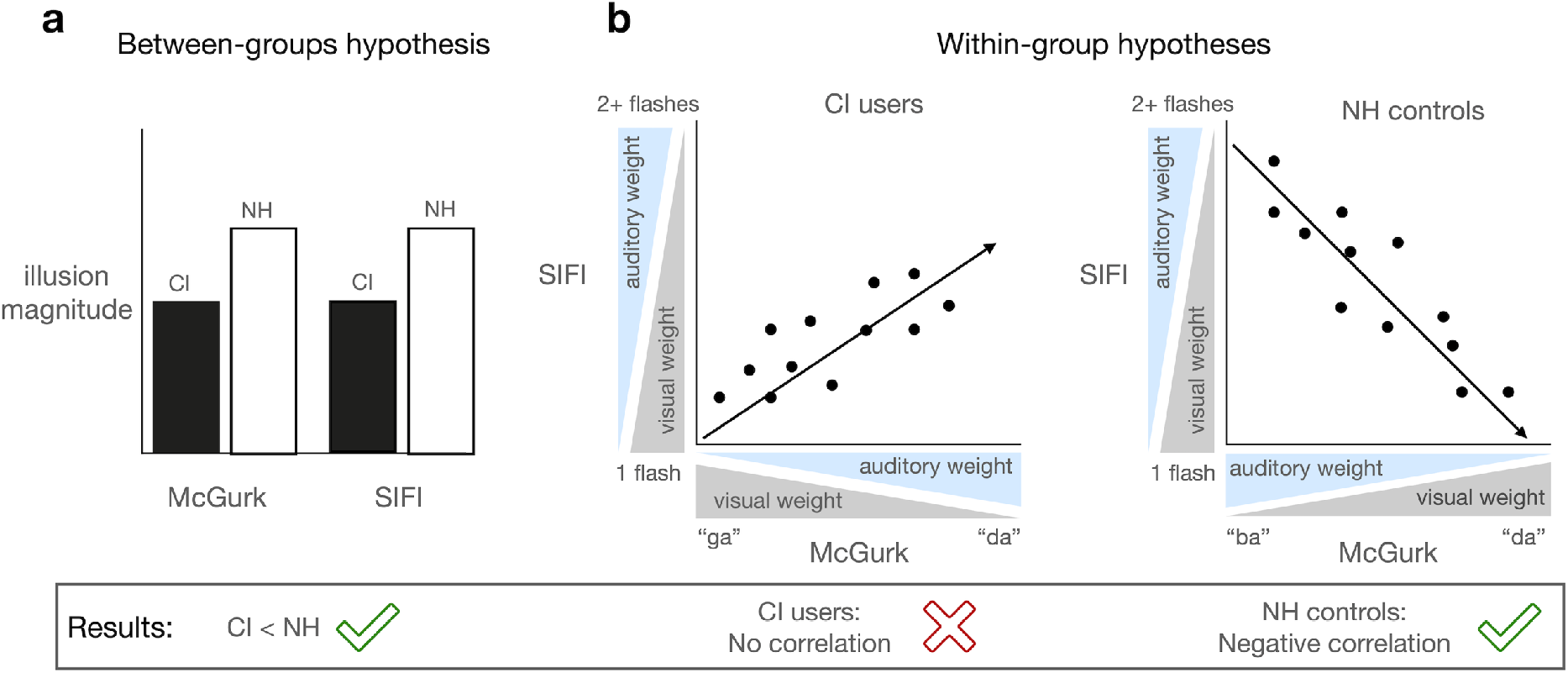
Schematic of main hypotheses and results. Data from this study supported our hypothesis that CI users experience the McGurk and Sound-Induced flash illusions less often than controls (a). We did not find a correlation between these two illusions in CI users as we expected (b), nor did they predict clinical speech outcomes beyond known metrics in the CI group (not pictured). We did, however, find a negative correlation between both tasks among the NH control group in support of our reasoning that their higher perceptual “weighting” of auditory information results in perceiving more illusory flashes (b).

This study tests the McGurk illusion in the largest CI cohort to-date and is the first to investigate the sound-induced flash illusion (SIFI) in this population. Our aim is to investigate how these tasks relate both to one another and to clinical outcome measures for CI users. Such work is necessary to characterize AV integration more completely in this cohort of individuals for whom it may be exceedingly important to “bind” auditory and visual information into more reliable multisensory percepts.

Our results indicate that CI users perceived both of these illusions less often than their normal-hearing counterparts. Within the CI group we did not find a correlation between the two illusions; however, NH controls do exhibit a weak negative correlation such that lower McGurk fusion (i.e., more auditory-dominant percepts) corresponds with more illusory flashes (SIFI). Given the absence of a correlation in the CI group, it is also possible that these two tasks may be mediated by different mechanisms of multisensory (i.e., audiovisual) integration that are disproportionally influenced by other demographic or clinical factors in CI users. Furthermore, a lack of predictive ability of these illusions in explaining clinical variability within the CI group highlights the need for more extensive clinical characterization that captures audiovisual processing proficiency among a population of individuals who critically rely on audiovisual integration for speech comprehension.

## METHODS

### Participants

We recruited 63 CI users and 69 NH controls (Table 2). There was no significant difference between groups for age (t_(130)_ = 1.9, p = 0.06, d = 20.6). At least 3 months of experience with their implants was an inclusion criterion for CI users, and the average was 4.6 ± 3.7 years post activation (range = 3 mo – 14 y). The duration of hearing loss as defined from the known onset was 23.6 ± 17.4 years on average (range = 1.5-75 y). We also calculated an approximate “duration of deafness,” which we defined as the date of cochlear implantation minus the onset of severe hearing loss. We used a standard definition of severe hearing loss as either the date when pure tone detection exceeded 70 dB HL or else the closest estimate from clinical records. The average duration of deafness was 2.4 ± 4.2 years (range 0 – 27 y). The majority of CI users (n = 53) were postlingually deafened; however, a subset was prelingually deafened (n = 10). These individuals received their implants at an average age of 3.5 ± 1.8 years (range = 1.3-6.5y; see black circles in Fig. 2). Although prelingual deafness (i.e., occurring prior to language acquisition and during critical periods of development) can negatively impact speech outcomes (Fitzpatrick 2015; Miyamoto et al. 1994), all of these 10 individuals achieved high speech proficiency (Fig. 2).

**TABLE 2.** Participant characterization. A large cohort of cochlear implant (CI) and normal hearing (NH) listeners ranging in age from 6 to 77 years old participated in this study. Nearly half of the CI users (48%) had bilateral implants while many of the others had residual acoustic hearing (41%) in the non-implanted ear. Among the 93 implanted ears in this study, the majority (60%) were manufactured by Cochlear, Ltd, followed by Med-El and Advanced Bionics.

| Group | N | Sex<br>(% female) | Mean age<br>± SD (y) | Number of<br>CIs | Acoustic<br>hearing | Implant<br>manufacturer | Duration of<br>hearing loss<br>(y) |
| --- | --- | --- | --- | --- | --- | --- | --- |
| CI | 63 | 56% | 44.6± 21.1 | one – 52%<br>two – 48% | 41% | 60% Cochlear<br>25% MED-EL<br>15% AB | 23.6 ± 17.4 |
| NH | 69 | 73% | 37.8± 20.1 | — | 100% | — | — |

**FIGURE 2.**
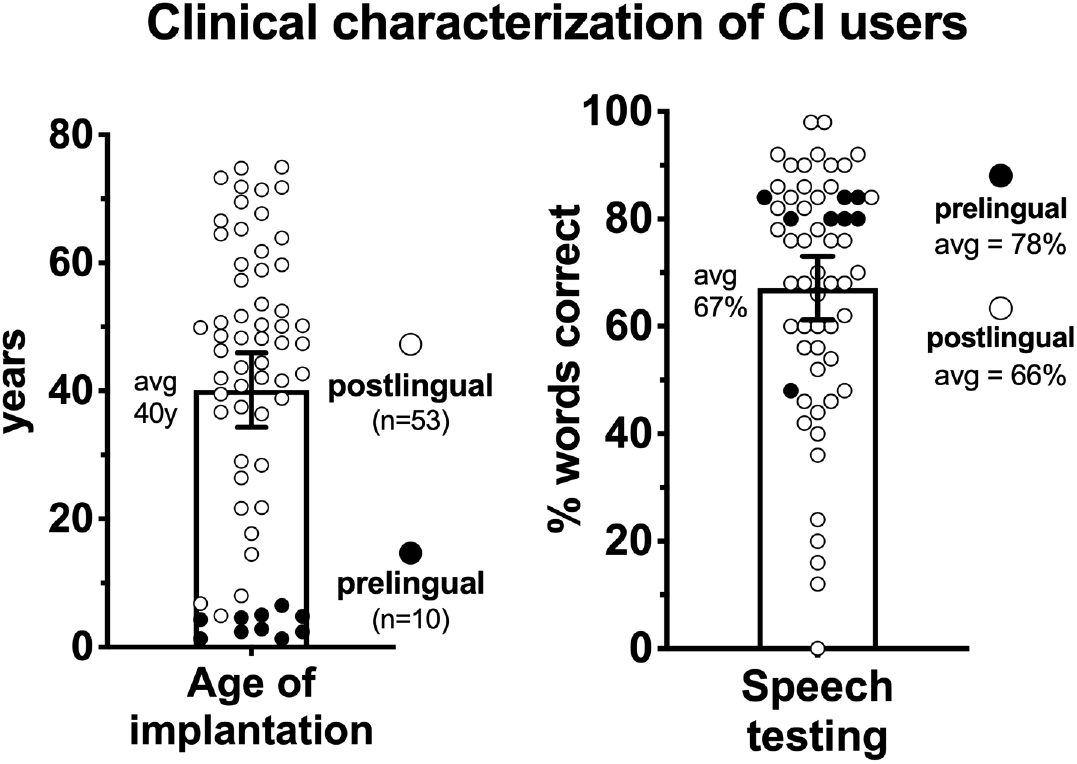
Clinical outcomes for the CI group. Age of implantation ranged from 1.3-75 years old (left), and all users had at least 3 months of CI use prior to participation. Clinical speech testing had a wide range of outcomes (right). We see higher than average performance in prelingually deafened individuals (filled circles) who were all implanted very young. Bars indicate group means and error bars are 95% confidence intervals of the mean.

All CI users completed testing in their “best-aided” hearing condition, which included hearing aids in the non-implanted ear for 41% of the sample. All participants wore corrective lenses as needed and were screened for visual acuity using either a Snellen eye chart or verbal report. Speech perception in the CI group was either tested at the study visit or recorded from recent medical records from the last 6 months. This measure was not available for four individuals, and of the remaining 59 CI users testing was carried out via best-aided, monosyllabic CNC word lists scored out of a possible 100% correct. Results indicate a wide range of proficiency from 0%-98% correct (67.1% ± 22.7%) (Fig. 2), and this variability is consistent with other reports in the literature (Gifford et al. 2018).

### Stimuli

Visual stimuli were displayed using Matlab 2008a and Psychophysics toolbox extensions (Brainard 1997). These stimuli were presented on a CRT monitor positioned approximately 50cm from participants. Visual stimuli were white circles (13 ms in duration at a 13° visual angle) on a black background and 2s videos of a female articulating the syllables “ba” and “ga” (Stevenson et al. 2014). Auditory stimuli were 3.5 kHz tones (50 ms in duration) in the SIFI task and utterance of the syllables “ba” and “ga” in the McGurk task. These auditory stimuli were delivered at a comfortably loud level (approximately 65 dB SPL) through a mono speaker. The aligned onset of visual and auditory stimuli was confirmed using a Hameg 507 oscilloscope via inputs from a photovoltaic cell and a microphone.

### Experimental Design and Analysis

The McGurk task has two blocks. The first block is unisensory testing of auditory-only and visual-only presentations of “ba” and “ga” (28 trials total). The next block is all audiovisual presentations of both syllables (20 trials each) and the incongruent pairing of auditory “ba” with the visual articulations of “ga” (20 trials). Participants responded to each trial with one of four letters corresponding to the sounds: “ba”, “ga”, “da”, or “tha” (Fig. 3). Illusory responses were considered either “da” or “tha”, which we will simply refer to as “da” throughout. Because misidentifying unisensory stimuli as one of these novel syllables could overinflate the apparent magnitude of the illusion, we made a correction using the formula:

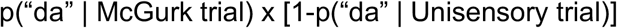

**FIGURE 3.**
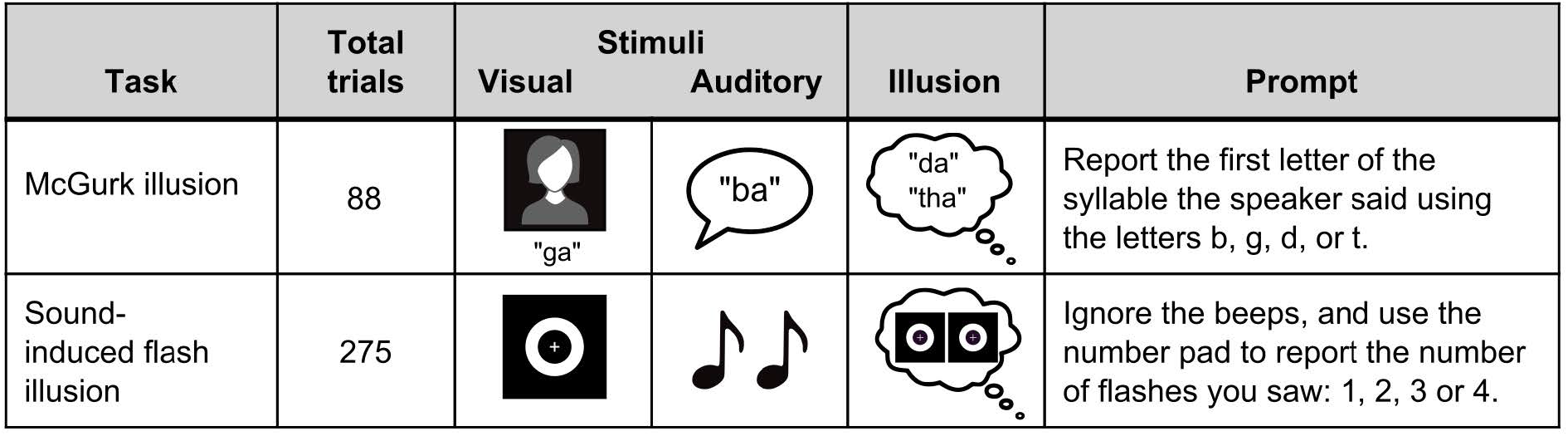
Experiment details of the McGurk illusion and the sound-induced flash illusion (SIFI). These two tasks have both congruent control trials and incongruent illusory trials where participants are asked to either report what the woman said (McGurk) or the number of flashes that they saw (SIFI).

If a participant perceives a unisensory component as “da”, that obfuscates whether a subsequent audiovisual perception of “da” is illusory and thereby indicative of integration. This formula controls for those occasions by subtracting “da” responses in unisensory trials from “da” responses to McGurk trials, which effectively lowers the probability of experiencing the illusion —p(McGurk)—for those who struggle to distinguish the component syllables on their own. In the SIFI task, participants fixated on a white cross in the middle of the screen while a white ring flashed on a back background (Fig. 3). They were asked to report the number of flashes while ignoring the beeps. Control trials consisted of: 1-4 flashes without beeps, congruent numbers of flashes and beeps (up to 4), and incongruent pairings of just one flash with 2-4 beeps. 25 trials were tested for each of these 11 conditions, and responses were scored as the average number of flashes reported. Additionally, we calculated a susceptibility index (SI; Stevenson, Zemtsov, and Wallace 2012) using the following formula where R_n_ is the average number of flashes for n beeps:

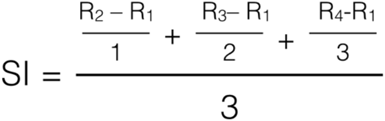

This index collapses across all incongruent trials in order to derive a single metric for the average number of illusory flashes that are experienced per added beep. A value of 0 means that no illusion was perceived, and a value of 1, for instance, indicates that each additional beep increased the perceived number of flashes by 1.

### Procedures

All protocols and procedures were approved by Vanderbilt University Medical Center’s Institutional Review Board, and all subjects provided informed consent prior to participation. Experiments took place in a dimly lit, sound-attenuated room with an experimenter seated nearby. Both task order and trial order were pseudo randomized. On all non-speech tasks, subjects were instructed to maintain fixation on the centrally-located fixation cross. All responses were collected using a standard keyboard. These experiments were part of a larger testing battery in which both trial and task order was pseudorandomized (Butera et al. 2018). Because some individuals ran out of time at the end of testing, subject numbers are included on figures to indicate those who were able to complete each task. In the McGurk experiment, one CI user and 2 NH controls did not have sufficient time to complete the unisensory testing necessary for p(McGurk) calculations. In lieu of imputation, we used the proportion of “da” responses as a proxy for p(McGurk) in these three individuals.

### Statistical approach

Between-group differences in the McGurk task were tested using resampling methods on account of several highly skewed variables that are incompatible with parametric tests even after standard transformations (e.g., log_10_). This approach was selected for its minimal assumptions of the data’s distribution in each group, and is based on 30,000 reshuffles where each shuffle reassigns data points randomly to the two groups. A two-tailed, Welch’s t-test is then used as a comparison metric. We selected this metric instead of a difference in means, for instance, because it has the advantage of capturing both a central tendency and variability. Significance is expressed as the number of times a shuffled sample produces a p value exceeding what was found in the actual sample, and is expressed as a proportion of the total number of simulations. T scores and degrees of freedom are reported from the Welch’s t-test of the observed data. Control McGurk trials, the p(McGurk) index, and SI were compared between groups using this resampling method in R (R Core team 2013).

Data from the SIFI task were less skewed and a square root transformation corrected for small deviations from normality. Congruent and incongruent conditions were both compared via a mixed-model ANOVA in SPSS. The repeated measure is the number of added beeps, and the two groups are the between-subjects factor. Greenhouse-Geisser corrections for violations of sphericity are applied as needed, and significant interactions are followed up with pairwise t-tests.

Correlations between tasks metrics were done with a Spearman’s rank-order correlation in SPSS software. Lastly, for a linear regression within the CI group, we included uncorrelated clinical variables with log transformations as needed to correct for deviations from normality.

Because prelingual deafness and subsequent pediatric implantation is frequently considered to be a distinct subgroup of CI users with a unique developmental time course, we also conducted all analyses excluding data from these 10 individuals. Since none of the main findings were affected by their inclusion, all statistical results and plots include all data collected, including both pre and postlingually deafened CI users. Significance for all statistical tests was defined as α < 0.05, and throughout, effect sizes are reported where applicable and means are reported ± standard deviations. De-identified data from this study are available from the corresponding author (I.M.B.) upon reasonable request.

## RESULTS

### McGurk Illusion

In both the unisensory and AV control (i.e., congruent) trials, CI users have lower speech perception accuracy for both “ba” and “ga” stimuli (Table 3). Not surprisingly, the group differences are largest for the auditory-only conditions (Fig. 4a). Additionally, for identifying “ga”, CI users had lower accuracy in lipreading (CI mean= 53%, NH mean = 68%) and AV listening (CI mean= 83%, NH mean = 99%). Though AV identification of “ba” was also statistically lower for CI users, both groups scored close to ceiling (CI mean =97.7% and NH mean= 99.3%).

**TABLE 3.** McGurk results. Control trials and incongruent AV “McGurk” trials are compared between groups and values (p< 0.05) are bolded.

| Modality | Syllable | t statistic | Significance | Effect size | TABLE 3. McGurk results. |
| --- | --- | --- | --- | --- | --- |
| Auditory-only | Ba | $t_{(92.2)} = -9.6$ | **p < 0.001** | d = 1.72 | Control trials and<br>incongruent AV “McGurk”<br>trials are compared<br>between groups and values<br>(p< 0.05) are bolded. |
| | Ga | $t_{(62.3)} = -6.8$ | **p < 0.001** | d = 1.23 | |
| Visual-only | Ba | $t_{(94.3)} = -1.4$ | p = 0.16 | d = 0.25 | |
| | Ga | $t_{(125.9)} = -3.0$ | **p = 0.004** | d = 0.54 | |
| Audiovisual<br>(congruent) | Ba | $t_{(74.1)} = -2.3$ | **p = 0.02** | d = 0.42 | |
| | Ga | $t_{(63.4)} = -6.7$ | **p < 0.001** | d = 1.20 | |
| Audiovisual<br>(McGurk) | Ba | $t_{(70.5)} = -5.9$ | **p < 0.001** | d = 1.01 | |
| | Ga | $t_{(119.4)} = 6.0$ | **p < 0.001** | d = 1.05 | |
| | Da | $t_{(121.0)} = 0.15$ | p = 0.87 | d = 0.03 | |

**FIGURE 4.**
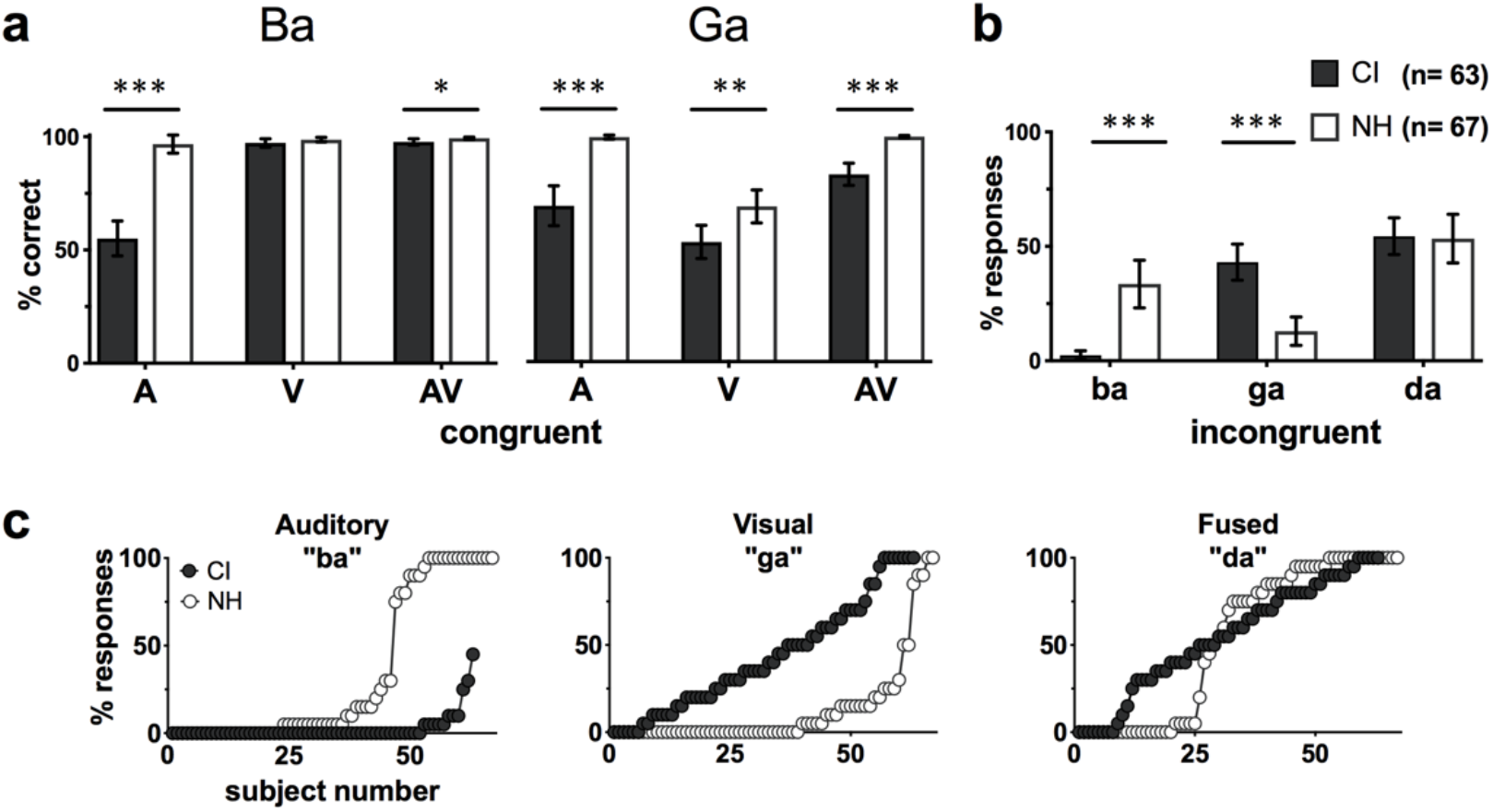
McGurk experiment results. (a) Mean accuracy of perceiving syllables in both unisensory and congruent AV trials are shown for “ba” and “ga” (a). (b) Mean responses to incongruent “McGurk” trials. (c) Individual data for each response type. Error bars indicate 95% confidence interval of the mean. * p< 0.05, **p< 0.01, ***p< 0.001.

In the incongruent AV (i.e., McGurk) trials (Fig. 4b), there is no significant difference in “da” responses (Table 3). However, when CI users did not fuse the syllables, they were much more likely to report the visual component “ga”. Conversely, non-fusing NH controls were most likely to report the auditory component “ba”. Individual data from these McGurk trials illustrate these biases as well as the high proportion of NH controls who did not perceive the illusion (Fig. 4c, white circles at or near 0% responses). In NH populations, prior studies have described the McGurk illusion occurring as an “all-or-nothing” effect such that the majority of individuals either experience the illusion almost always or else rarely ever (Mallick et al. 2015). In the present study, we found that 72% of the NH group fell within the two extremes of perceiving the illusion either ≥90% of the time or ≤10%. In contrast, the CI group had many more intermediate responses with only 35% of individuals having very high or very low fused responses (see “da” panel in Fig. 4c).

### Sound-induced flash illusion (SIFI)

In control trials where the number of flashes and beeps is matched (Fig. 5a), there are no between-group differences in the perceived flashes (F_(3,127)_ =0.002, p = 0.97, η_partial^2^_= 0). In the incongruent pairings of just one flash with multiple beeps (Fig. 5b), there is also no between-subjects effect (F_(4,127)_ =0.55, p = 0.46, η_partial^2^_= 0.004). However, there is a beep × group interaction (F_(1.4,127)_ = 7.6, p = 0.003, η_partial^2^_= 0.056). Follow-up tests did not identify any group differences at specific conditions, though several approached significance (Table 4). Most notably, CI users reported slightly fewer flashes in the three-beep (avg flashes = 1.96) and four-beep conditions (avg flashes = 2.07) compared to controls (2.16 and 2.30, respectively).

**TABLE 4.**
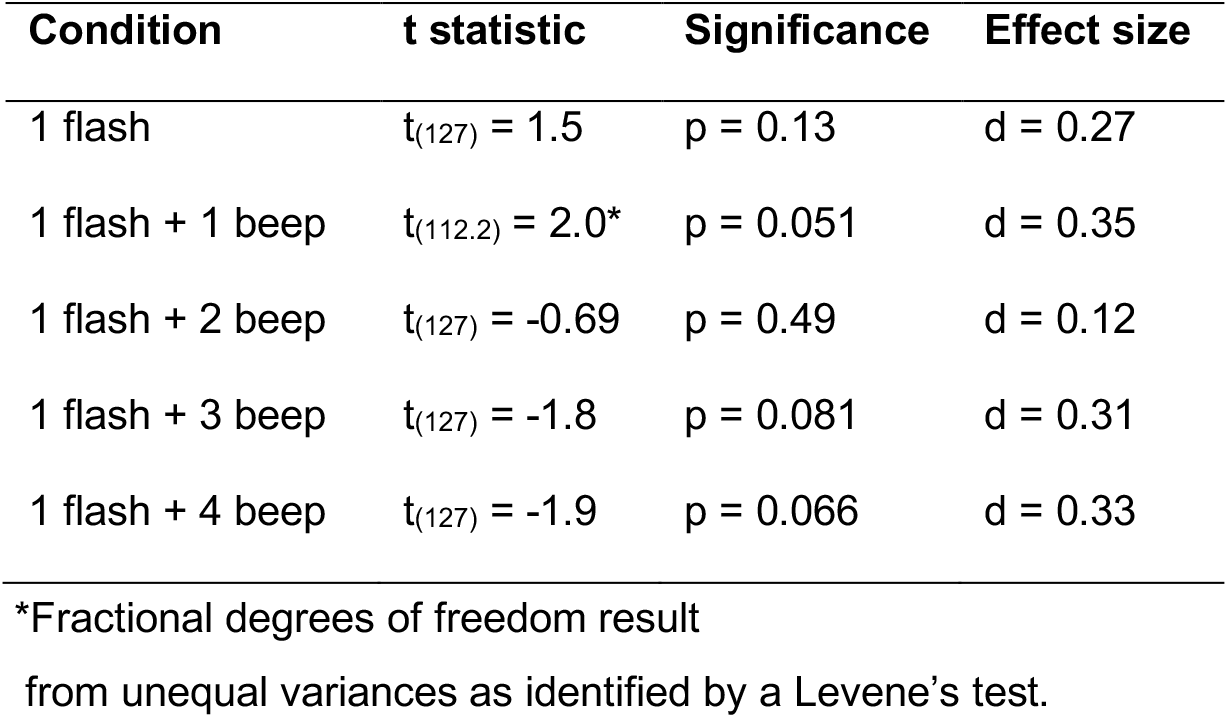
SIFI results. Though within-subjects effects from a mixed-model ANOVA indicated a significant condition × group interaction, no follow-up t-tests reached the p = 0.05 significance threshold.

**FIGURE 5.**
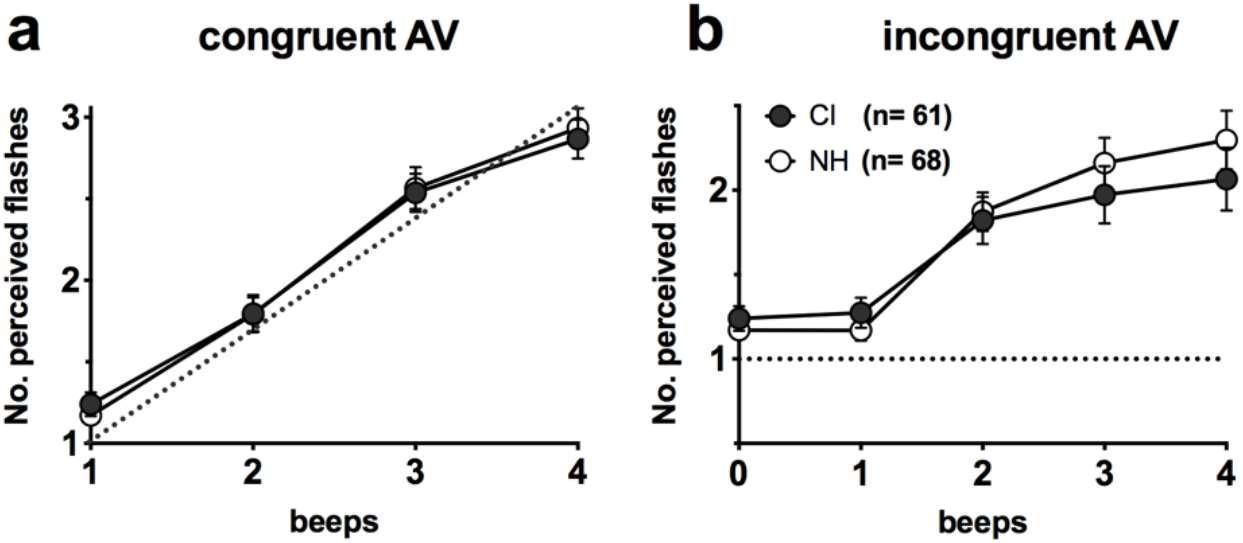
Sound-induced flash Illusion results. Average reports of the number of perceived flashes are plotted for each group. Trials were either congruent pairings of one or more flashes *and* beeps (a) or 0-4 beeps paired with only one flash (b). Dotted lines represent 100% accuracy and error bars are 95% confidence intervals of the mean.

### Illusion metrics

In order to compare group differences in illusion perception between these two tasks, we calculated p(McGurk) and SI metrics (Fig. 6). In an initial comparison of p(McGurk) between groups, there is no significant difference (t_(109.6)_ = −1.7, p =0.097, d= 0.33). However, we also compared whether the magnitude of the illusion differed strictly among individuals who did perceive the illusion in at least one trial. Thus, we excluded all “non-perceivers” who made up 13% of the CI group (n = 8) and 30% of the NH (n = 20). Of the remaining 55 CI users and 47 NH controls, we see a highly significant difference between p(McGurk) measures (t_(81.0)_ = −4.9, p = 4×10^−5^, d= 0.24). Following the same trend, there is also significantly lower SIFI susceptibility for CI users (t_(124.5)_= −2.5,p = 0.014, d= 0.31; Fig. 6b).

**FIGURE 6.**
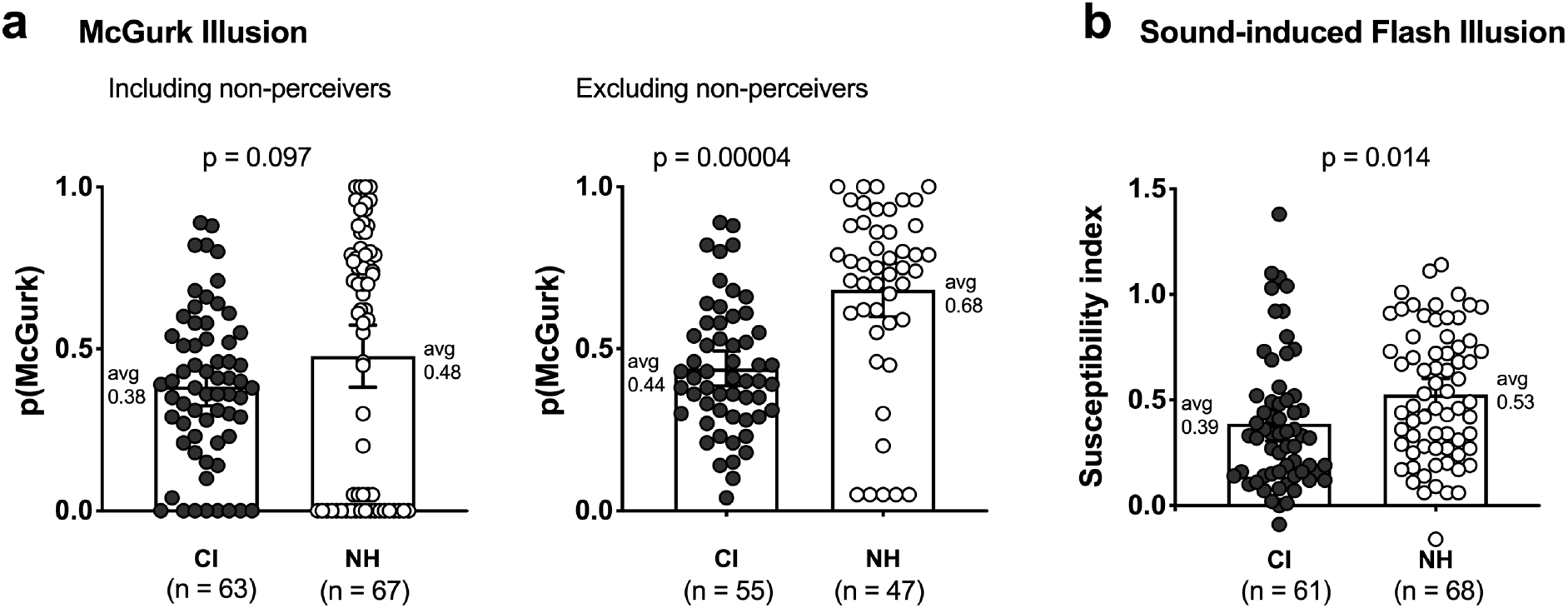
Derived metrics of the probability of McGurk perception and SIFI susceptibility index. The p(McGurk) metric corrects for inaccurate “da” percepts in control conditions (i.e., which only contain “ba” and “ga”). Susceptibility index collapses across all incongruent conditions (i.e., 2 to 4 beeps per flash) to quantify the average increase in perceived flashes per added beep.

### Correlation between illusion tasks

Next, we asked whether these two illusions have a relationship to one another in these cohorts. We found no correlation between the illusions for the 87% of CI users who experienced the McGurk effect (r_s_ = 0.041, p=0.77, Fig. 6a); however, we did find a negative, albeit weak, correlation among the 70% of NH controls who perceived the McGurk illusion (r_s_ = −0.317, p=0.030).

### Testing for additional explained variability in clinical measures

Lastly, we asked whether these AV illusion metrics have any predictive value for explaining variability in clinical speech scores within the CI group (Fig. 2). In a stepwise regression model with CNC scores as the dependent variable, we entered the following four uncorrelated clinical and experimental measures as independent variables: duration of hearing loss, duration of deafness, p(McGurk), and SI. The only variable that is a significant predictor of CNC scores is the duration of hearing loss, explaining 13% of variability (R^2^ = 0.130, F_(1,40)_= 5.97, p= 0.019). All other variables were excluded from the model (Table 5).

**TABLE 5.**
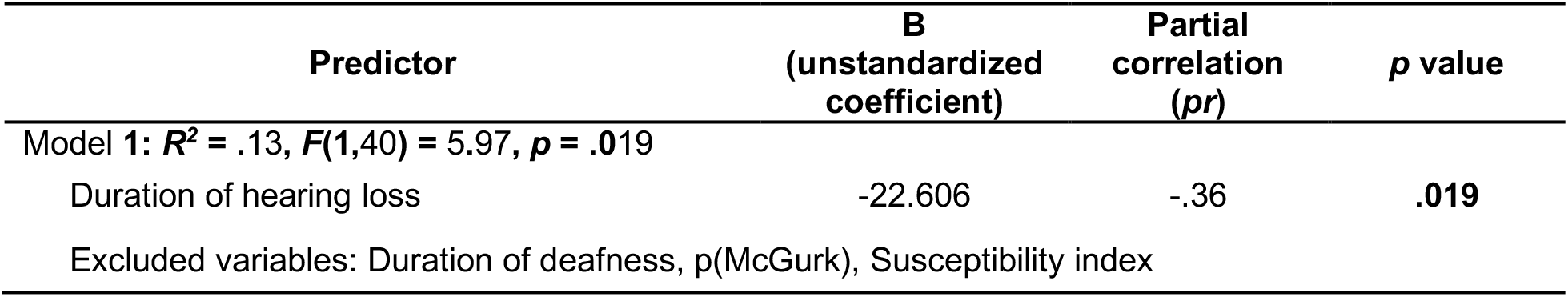
Predictors of CNC variability. In a stepwise linear regression with CNC speech scores as the dependent variable, illusions metrics did not significantly contribute to the model beyond the 13% of variability predicted by duration of hearing loss.

## DISCUSSION

This study tested the McGurk illusion in the largest CI cohort to-date and is the first to investigate the sound-induced flash illusion in this population. A key finding is that CI users perceived both of these AV illusions less often than NH controls (Fig. 6). Additionally, we replicated the same general trend as others have reported in the McGurk task: that CI users display a bias toward the visual speech syllable (Fig. 4). Importantly, our results also illustrate the need to correct for mistaking unisensory syllables as “da”. That is, in this study, like several others (Huyse et al. 2013; Rouger et al. 2008; Tremblay et al. 2010), raw data of “da” responses were the same between groups (Fig. 4c). It was only in comparing the p(McGurk) measure of individuals who perceived the illusion at least once that we saw the groups significantly diverge (Fig. 6). Based on these findings, we recommend that any further investigations of this illusion in CI users take similar measures to disambiguate “da” responses and fused percepts to avoid overestimating the magnitude of the McGurk effect.

Similarly, in the SIFI task, CI users were able to perceive the illusion; however, the magnitude of the effect was lower than for controls. We saw small deviations in several flash-to-beep ratios (Table 4) that, after collapsing across all conditions into a single susceptibility metric, was significantly lower than the control group (Fig. 6b). This novel finding is noteworthy given that relatively little is known about low-level audiovisual stimulus detection in CI users (Stevenson et al. 2017). However, based on our prior work, we do know that CI users’ temporal judgments of synchrony for these same flashbeep stimuli are indistinguishable from controls in a simultaneity judgement experiment (Butera et al. 2018). This suggests that low-level AV temporal function is likely intact in adult CI users, so any broader issues may be subtle. Although work in other clinical populations has related reduced SIFI perception to broader impairments in integration (Stevenson et al. 2014), additional work is needed to better characterize any further implications and mechanisms in CI users.

A broad interpretation of the present study is that CI users perceptually “weight” visual information more highly, which has a consequent impact on both the McGurk and SIFI illusions. That is, in incongruent tasks where illusory percepts are more heavily dependent upon sound biasing the visual cue, CI users are more likely to have strictly-visual percepts. This visual bias translates to fewer illusory flashes in SIFI and more frequent visual “ga” percepts instead of fusion with auditory information in the McGurk illusion. A similar pattern of more visually-biased McGurk responses can also be simulated in NH users by vocoding stimuli to sound like signals derived from a cochlear implant (Desai et al. 2008). Similarly, adding a visual distractor reduces McGurk percepts (Tiippana, Andersen, and Sams 2004), as does adding noise to reduce the auditory and/or video salience (Fixmer and Hawkins 1998; Hirst et al. 2018).

Further work is needed to understand the clinical impacts of this visual bias in CI users. Though prior studies have suggested that more proficient CI users experience greater fusion (and less visual bias) (Huyse et al. 2013; Tremblay et al. 2010), we did not find McGurk fusion to explain any greater variability in CNC scores than other known measures—in our case, the total duration of hearing loss (Table 5). A major caveat to consider in interpreting these findings is that our outcome measure, which is standard in clinical practice, is auditory only. As a result, it is perhaps unsurprising that an audiovisual illusion like the McGurk effect may relate more directly to audiovisual speech measures than auditory-only ones (i.e., CNC scores). However, as others have reported in NH samples, correlations between McGurk results and audiovisual speech measures are weak (Van Engen, Xie, and Chandrasekaran 2017), and it is likely that many studies are underpowered to detect such relationships (Magnotti et al. 2020). As a result, larger studies may be necessary to draw the presumed connection between McGurk susceptibility and other AV speech measures, particularly those involving natural, congruent speech.

Though the stimuli in the McGurk and SIFI tasks are dissimilar, perceiving both of these AV illusions may have shared mechanisms since both require the viewer to reconcile whether incongruent auditory and visual cues do in fact originate from the same source (Körding et al. 2007). When comparing the illusion magnitudes between these two experiments, we found a negative correlation between the SI and p(McGurk) metrics in the NH group (r_s_ = −0.317, p = 0.030). However, we did not see a direct relationship between these measures in CI users (Fig. 7) as we had anticipated (Fig. 1b). In a slightly younger and smaller NH sample size, Stevenson and colleagues (Stevenson et al. 2012) reported a stronger correlation between p(McGurk) and SIFI (R^2^=0.42 p = 0.003) that also had a negative relationship. In contrast, Tremblay et al. (2007) found no correlation between SIFI and McGurk tasks in a younger sample of 38 typical listeners between 5 and 19 years of age. While it is logical that differences in low-level AV illusions indexed by SIFI could have cascading effects on speech integration, these two tasks do not appear to have a direct relationship in CI users. Further work to model the process of cue combination in each of these tasks may provide additional context for how subject-specific parameters like sensory noise may correspond between McGurk and SIFI experiments (Magnotti and Beauchamp 2015; Magnotti, Ma, and Beauchamp 2013).

**FIGURE 7.**
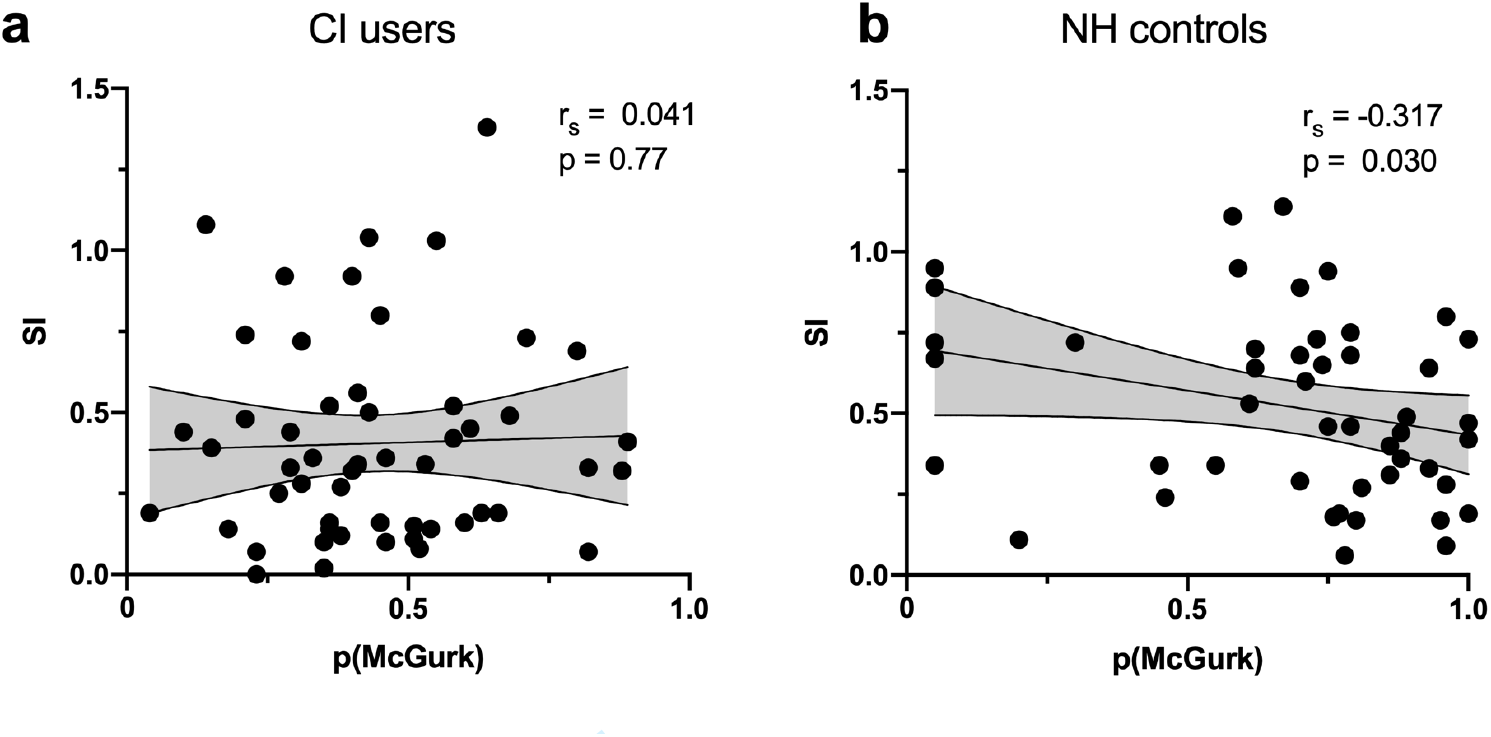
Relationship between SIFI and McGurk. The correlation between susceptibility index (SI) and probability of McGurk perception is not significant for CI users (a), but does have a significant negative relationship among NH controls (b). Shaded areas correspond to 95% confidence intervals.

It should be noted that the illusion of multiple flashes can be elicited in other modalities as well (Violentyev, Shimojo, and Shams 2005). In an fMRI study of deaf individuals, a visual-tactile illusion that is analogous to SIFI, revealed larger visual responses in Heschl’s gyrus and a greater magnitude of the illusion experienced in deafness (Karns, Dow, and Neville 2012). Although it’s beyond the scope of the present study, it would be interesting for future work to investigate possible underlying mechanisms. For instance, given what is already known about the mechanisms behind the sound-induced flash illusion (Cecere, Rees, and Romei 2015; Kerlin and Shapiro 2015; Watkins et al. 2006), it would be interesting to test whether, for instance, visually-derived alpha oscillations play a similar role in predicting the likelihood of experiencing the illusion in CI users or if other, crossmodal mechanisms are at play (Stropahl and Debener 2017).

One caveat to our study is that that McGurk perception is highly stimulus-specific, and in our case, we only tested responses to a single female speaker. As a result, it is possible that these results may not generalize more broadly to other syllable combinations, male speakers, languages, etc. The primary advantage for interpreting our findings, however, is that we tested a large sample of clinically-diverse CI users, so CI perception to this particular stimulus is well-characterized. In future studies, applying a noisy encoding of disparity (NED) model to quantify results would be ideal for investigating the effect in a stimulus-independent manner (Magnotti and Beauchamp 2015; Stropahl et al. 2017). Such work may also provide more context for our curious findings that CI users have lower lipreading proficiency with the syllable “ga,” as well as lower identification of the congruent, audiovisual presentation of “ga” (Fig. 4a). In the past our lab has recorded and assayed several other McGurk stimuli and have continuously found the present stimuli to elicit the largest magnitude of the McGurk effect (unpublished observation).

This may be due to a high ambiguity of the visual component in this particular recording, which could have disproportionately obfuscated the speech content for proficient lip readers like CI users. Thus, CI users performed more poorly in these control conditions for the same reason that this is a good McGurk stimulus—the articulation of “ga” (as well as the speaker’s pre-articulatory movements) is particularly ambiguous and more like “da” than other speakers/recordings. Again, the methods described by Magnotti and colleagues (2015) would address this issue more conclusively.

Finally, it is not entirely clear what these results mean for cochlear implant outcomes given the lack of a relationship with CNC scores. Are more visually-biased CI users less proficient with their implants? Are they worse “multisensory integrators” on broader—and more practical—AV speech tasks or simply less apt to perceive illusions? Because it’s more likely that these AV illusions would relate to audiovisual speech measures than the auditory-only ones tested in clinic (and reported here), future work comparing illusion perception to a real-world estimate of one’s success in the integration of conversational, audiovisual speech would provide more insight. Until we know how McGurk biases relate to natural speech integration of words or sentences it is unclear what utility administering McGurk testing at a broader scale might have. Either way, better identifying individual differences in multisensory integration is an important next step, particularly for the development of new audiovisual remediation strategies.

## Data Availability

De-identified data from this study are available from the corresponding author (I.M.B.) upon reasonable request.

## ACKNOWLEDGEMENTS

We would like to thank the volunteers who participated in this study, and the assistance provided by Brannon Mangus and Amelia Schuster with recruitment as well as the statistical guidance provided by Dan Ashmead.

